# Wuhan and Hubei COVID-19 mortality analysis reveals the critical role of timely supply of medical resources

**DOI:** 10.1101/2020.03.13.20035410

**Authors:** Zuqin Zhang, Wei Yao, Yan Wang, Cheng Long, Xinmiao Fu

**Author notes:** To whom correspondence should be addressed to Professors Xinmiao Fu and Dr. Cheng Long. These authors contributed to this work equally (Zuqin Zhang, Wei Yao).

## Abstract

We report that COVID-19 mortality and recovery rates in Hubei Province, China exponentially decays (*R*^*2*^>0.93) and grows (*R*^*2*^>0.95), respectively. A great number of newly supplied medical resources, including more than 42000 aided health workers, over 26000 makeshift beds and 23000 acute care beds, enabled overwhelming patients to be treated effectively in hospitals. This may inform other countries to deal with the coming COVID-19 pandemic when patients are overwhelming the local health care system.

---

The 2019 novel coronavirus diseases (COVID-19) outbreak caused by SARS-CoV-2 is on-going in China and has hit many countries ^1-3^. As of 3 March 2020, there have been 80270 confirmed cases and 2981 deaths in China, most of which are from the epicenter of the outbreak, Wuhan City, the capital of Hubei Province. New COVID-19 cases have been steadily declining in China and more than 60000 patients have been recovered ^4^, largely due to the effective implementation of comprehensive control measures in China ^5, 6^. Here we report that some of these measures, such as a dramatic and timely increase of medical supplies, may play a critical role such that the mortality and recovery rates of COVID-19 in Wuhan follow exponential decay and growth modes, respectively.

We collected data for analysis on the officially released cumulative numbers of confirmed, dead and recovered cases (from 23 Jan to 3 Mar 2020) in five geographic regions, i.e., mainland China, Hubei Province, outside Hubei (in China), Wuhan City and outside Wuhan (in Hubei). As of 3 Mar 2020, crude fatality ratios (CFRs) in the above regions are 0.027±0.006, 0.035±0.007, 0.005±0.002, 0.045±0.012 and 0.021±0.008, respectively, in line with earlier reports ^5, 6^. While the mortality rates of COVID-19 outside Hubei and outside Wuhan appear constant over time, the mortality rates in Hubei and Wuhan decline continuously (**Fig. 1A**). Strikingly, the mortality rates in Hubei and Wuhan are well-fitted with the exponential decay mode (*R*^*2*^ being 0.93 and 0.82, respectively; **Fig. 1A** and **Table S1**), and it is the same forth with that in China (*R*^*2*^ being 0.86) but not with that outside Hubei and outside Wuhan (*R*^*2*^ being 0.39 and 0.32, respectively). Remarkably, we found that the recovery rates of COVID-19 patients in the above regions were all well-fitted with the exponential growth mode (*R*^*2*^ being 0.96, 0.95, 0.95, 0.88 and 0.95, respectively; **Fig. 1B** and **Table S1**). Such intriguing pattern for the COVID-19 mortality and recovery rates in Wuhan (or Hubei) somehow contradicts traditional epidemiological models wherein both are assumed as constants ^7^.

**Fig. 1.**
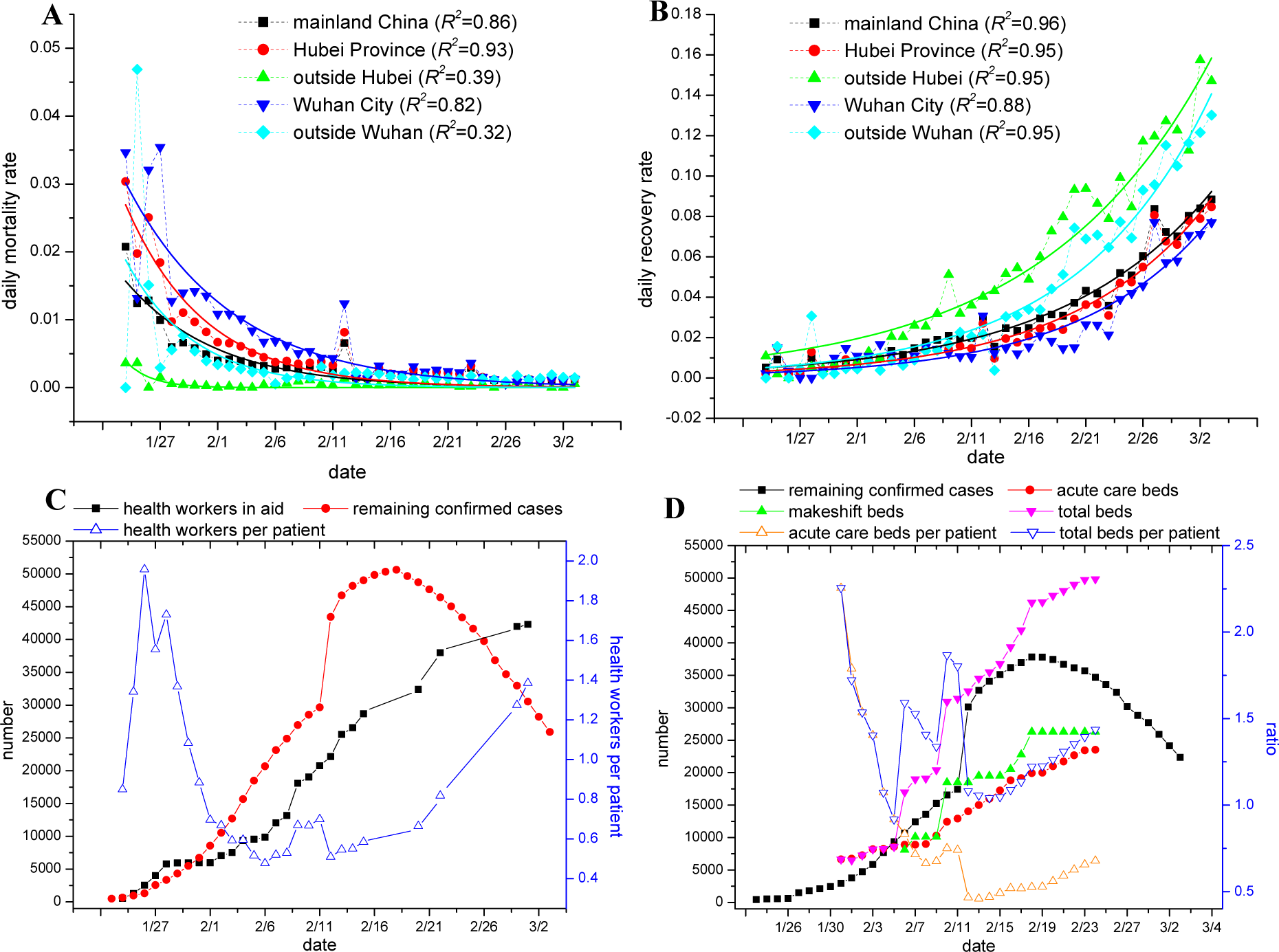
Fitting the COVID-19 mortality and recovery rates with exponential decay and growth functions, respectively. (**A, B**) Mortality rate (panel A) and recovery rate (panel B) for COVID-19 in China over time and by location, with exponential decay- and growth-based regression analyses being performed, respectively (as shown by colored solid lines). Parameters from the regression analyses are shown in **Table S1**, with *R*^*2*^ being shown here. (**C**) Numbers of the aided health workers in Hubei over time. Ratio of the aided health workers to patients was also plotted (note: most of the aided health workers are working in Wuhan; refer to **Table S3**). (**D**) Numbers of the remaining confirmed cases of COVID-19, and acute care beds, makeshift beds from Fangcang hospitals and total beds in Wuhan over time. Ratio of beds to patients was also plotted. Here the data of newly supplied beds in Hubei are not available and thus the data for Wuhan were analyzed.

The above unique pattern may reflect the fact that COVID-19 patients in Wuhan (or Hubei) have been treated more effectively day by day. Here we focused on two components essential for effective treatments, i.e., the supply of health workers and hospital beds. As a matter of fact, a great number (up to 42 000, as of 1 March 2020) of health workers have been aided by other provinces in China (**Table S2**) and they are working in different cities of Hubei (**Table S3**). This extraordinary aid keeps the ratio of the health workers to patients in Hubei at above 0.6 despite the number of remaining confirmed cases has ten-fold increased up to 50 000 on 18 Feb (**Fig. 1C**). Results also show that the number of acute care beds from more than 45 designated hospitals plus two newly built ones in Wuhan has been consecutively increasing up to 23 532 (as of Feb 24) under the government-directed re-allocation (**Fig. 1D**). This supply thus enabled the severe and critical patients to be treated timely and effectively ^6^. More importantly, there have been over 10 temporary hospitals (named Fangcang hospitals) reconstructed from gymnasium and exhibition centers, which provide more than 26 000 makeshift beds for mild patients (**Fig. 1D**). These combinations guaranteed nearly 100% percent of COVID-19 patients to be treated in hospitals even if the number of remaining confirmed cases has ten-fold increased up to 38 000 on 18 Feb (**Fig. 1D**). In contrast, a lot of patients had to stay at home in the early stage of the outbreak in Wuhan due to the shortage of beds such that many transmissions in households occurred ^6^.

Accordingly, the effective implementation of comprehensive control measures and infection-treatment practices is critical for combatting any new pathogens, not only interrupting the transmissions but also saving the patients. Timely supplied medical resources, including re-allocation of acute care beds, rapid construction of new hospitals and generous aid of health workers by other less-severe areas, apparently help the epicenter of the outbreak Hubei (Wuhan) to accomplish a unique and also encouraging outcome for life-saving such that the mortality and recovery rates of nearly 50 000 COVID-19 patients exponentially decays and grows, respectively. Other crucial factors contributing to this success may include the improved and optimized diagnosis and treatment strategies ^8^, which are critical for saving severe and critical patients ^5, 6, 9^. This speculation appears to be supported by the exponential growth of the COVID-19 recovery rate outside Hubei (**Fig. 1B**) where medical resources are relatively sufficient ^10^. Collectively, the achievement made in Hubei (or Wuhan) may provide useful guidance for many countries to be better prepared for the potential pandemic ^2^ that may overwhelm local health care systems.

## Data Availability

All the data are included in the manuscript

## Acknowledgments

This work is support by the National Natural Science Foundation of China (No. 31972918 and 31770830 to XF). All authors report no conflicts of interest relevant to this article.

